# Investigational treatments for COVID-19 may increase ventricular arrhythmia risk through drug interactions

**DOI:** 10.1101/2020.05.21.20109397

**Authors:** Meera Varshneya, Itziar Irurzun-Arana, Chiara Campana, Rafael Dariolli, Amy Gutierrez, Taylor K. Pullinger, Eric A. Sobie

## Abstract

Many drugs that have been proposed for treatment of COVID-19 are reported to cause cardiac adverse events, including ventricular arrhythmias. In order to properly weigh risks against potential benefits, particularly when decisions must be made quickly, mathematical modeling of both drug disposition and drug action can be useful for predicting patient response and making informed decisions. Here we explored the potential effects on cardiac electrophysiology of 4 drugs proposed to treat COVID-19: lopinavir, ritonavir, chloroquine, and azithromycin, as well as combination therapy involving these drugs. Our study combined simulations of pharmacokinetics (PK) with quantitative systems pharmacology (QSP) modeling of ventricular myocytes to predict potential cardiac adverse events caused by these treatments. Simulation results predicted that drug combinations can lead to greater cellular action potential prolongation, analogous to QT prolongation, compared with drugs given in isolation. The combination effect can result from both pharmacokinetic and pharmacodynamic drug interactions. Importantly, simulations of different patient groups predicted that females with pre-existing heart disease are especially susceptible to drug-induced arrhythmias, compared males with disease or healthy individuals of either sex. Overall, the results illustrate how PK and QSP modeling may be combined to more precisely predict cardiac risks of COVID-19 therapies.

## INTRODUCTION

The rapid spread of COVID-19 worldwide, in the absence of established therapeutics, has forced clinicians to improvise by treating patients “off label” with drugs that were approved to treat other diseases. The investigational off label use of such drugs for COVID-19 has led to the initiation of several clinical studies, generally small in scope due to the urgent nature of the health care crisis. Some of these have included drug combinations, such as lopinavir plus ritonavir^1^ or chloroquine plus azithromycin.^2^ Under the current pandemic conditions, with clinicians attempting to maintain scientific rigor while delivering treatments quickly, it can be difficult to properly weigh the potential benefits of drugs against the risks of adverse events. It can be especially challenging to determine the risks of drug combinations because specific combinations have often not been examined during preclinical or early clinical safety assessments.

Mathematical models can be used to rapidly predict the physiological effects of drug treatments, and simulations with such models gain outsize importance when time is of the essence. This can become especially true when considering drug combinations due to the complexities that are introduced when drugs are co-administered. These can include both pharmacokinetic (PK) interactions, whereby the presence of one drug alters the concentration of a second drug, and pharmacodynamic interactions, whereby the overall physiological consequences result from the combined biological effects of the two drugs. The former can be studied with pharmacometrics approaches, whereas the latter can be addressed with quantitative systems pharmacology (QSP) models that explicitly incorporate drug mechanisms of action. Although simulation results are always subject to model limitations, they can nonetheless guide decision making. Both PK and QSP simulations are likely to become particularly important for proposed COVID-19 treatments because: (1) adverse cardiac events, including QT prolongation, have been associated with several of these drugs,^3, 4, 5^ (2) drug disposition has previously been characterized for most of the drugs being considered for treatment;^6, 7, 8^ and (3) modeling of pharmacological effects on cardiac electrophysiology, including adverse events, is a mature area of research.^9, 10, 11^

Here we present cellular simulation results that predict an additive drug combination effect on QT prolongation, indicating that these combinations may increase the risk of ventricular arrhythmias in COVID-19 patients. The simulations further highlight the importance of sex differences and the presence of existing comorbidities such that particular drug combinations may be especially dangerous for women with heart failure. Importantly for future studies, the results demonstrate a pipeline for systematic examination and a quantitative methodology that can be used to balance the potential benefits of COVID-19 treatments against the risks of cardiac arrhythmias.

## METHODS

### Drugs considered and data sources

To begin to understand potential side effects caused by COVID-19 treatments, we considered 4 drugs currently under investigation: lopinavir, ritonavir, chloroquine, and azithromycin, as well as the former two and the latter two drugs in combination. We selected these 4 drugs for initial analysis because their effects on cardiac ionic currents have been assessed under standardized conditions. Crumb et al^12^ have reported effective free therapeutic plasma concentrations (EFTPCs) for each drug in addition to half maximal inhibitory concentration (IC_50_) values that indicate the affinity of each drug to block the following cardiac ionic currents: (1) late Na^+^ current (I_NaL_); (2) transient outward K^+^ current (I_to_); (3) rapid delayed rectifier K^+^ current (I_Kr_); (4) slow delayed rectifier K^+^ current (I_Ks_); (5) inward rectifier K^+^ current (I_K1_); (6) L-type Ca^2+^ current (I_CuL_). This unified data source was exploited for QSP model simulations of drug effects (see below).

### Cardiac cellular QSP modeling of drug effects

The O’Hara et al mathematical model^13^ of the human endocardial ventricular myocyte was used to simulate the effects of drugs on ventricular action potentials (APs). This model, comprising a system of 41 ordinary differential equations, simulates interactions between ionic currents and Ca^2+^ cycling in the ventricular myocyte, and drug-induced changes to APs in models such as these are well-correlated with clinically observed changes to electrocardiographic waveforms.^9, 10, 11^

Block of ionic currents by particular drugs was simulated with a pore block model. With this approach, the conductance (G) of each ionic current is scaled based on drug concentration ([C]) and the IC_50_ value describing how the drug blocks that current, i.e.

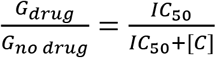

### Pharmacokinetic modeling

To link drug concentrations in the QSP simulations with free plasma drug concentrations that are likely to be observed in patients, we used PK models to simulate drug disposition of either azithromycin^8^ or lopinavir + ritonavir.^7^ These models, implemented as published, allowed us to simulate temporal changes in either total or free plasma concentrations with different dosing regimens. For lopinavir and ritonavir, we assumed an unbound fraction of 0.01 to relate total drug concentrations to free plasma concentrations that are likely to be present in patients.^14^ The azithromycin PK model^8^ calculated free drug concentrations directly. For chloroquine, which has been shown to accumulate in tissues including the heart, we extracted relevant cardiac drug concentrations from a physiologically-based PK model^6^ and assumed the unbound drug fraction in the heart (0.39) was identical to the fraction in plasma.^15^

### Pharmacodynamic variability between individuals

Two factors that are known to influence the risk of drug-induced arrhythmia are female sex^16^ and the presence of pre-existing heart failure (HF).^17^ We simulated differences between male and female and between healthy and failing myocytes using established protocols. These methods, published by Yang et al^18^ and Gomez et al,^19^ respectively, consist of scaling values for ionic current maximal conductances based on measured differences in ion transport pathways between male and female hearts, or between healthy and failing myocytes. Applying these scale factors allowed us to create 4 variants of the baseline ventricular myocyte: healthy male (assumed to be the original model), healthy female, HF male, and HF female. Once baseline models for the different patient groups had been created, we generated virtual populations to simulate physiological variability between individuals by randomly varying each baseline model’s maximal conductances, as previously described.^20, 21, 22^

## RESULTS AND DISCUSSION

We performed simulations to predict the effects on human ventricular action potentials (APs) of lopinavir, ritonavir, chloroquine, and azithromycin. Based on a comprehensive study of how various drugs affect cardiac ion channels,^12^ we calculated that each of the proposed COVID-19 treatments will block a slightly different complement of ionic currents (**Fig. 1A**). These drug-specific, concentration-dependent alterations to ionic currents were applied to a mathematical model of the human ventricular endocardial myocyte^13^ to predict drug-induced changes to cardiac APs. Simulations performed with each drug at 10 times the reported^12^ EFTPC show that all drugs can induce substantial AP prolongation at these high concentrations (**Fig. 1B**), with lopinavir causing the most dramatic effects. Simulations performed across drug concentrations ranging from 0.3 times to 10 times EFTPC confirm that the drugs prolong AP duration at 90% repolarization (APD_90_) in a concentration-dependent manner, with the largest effects occurring at the highest drug concentrations (**Fig. 1C**).

**Figure 1:**
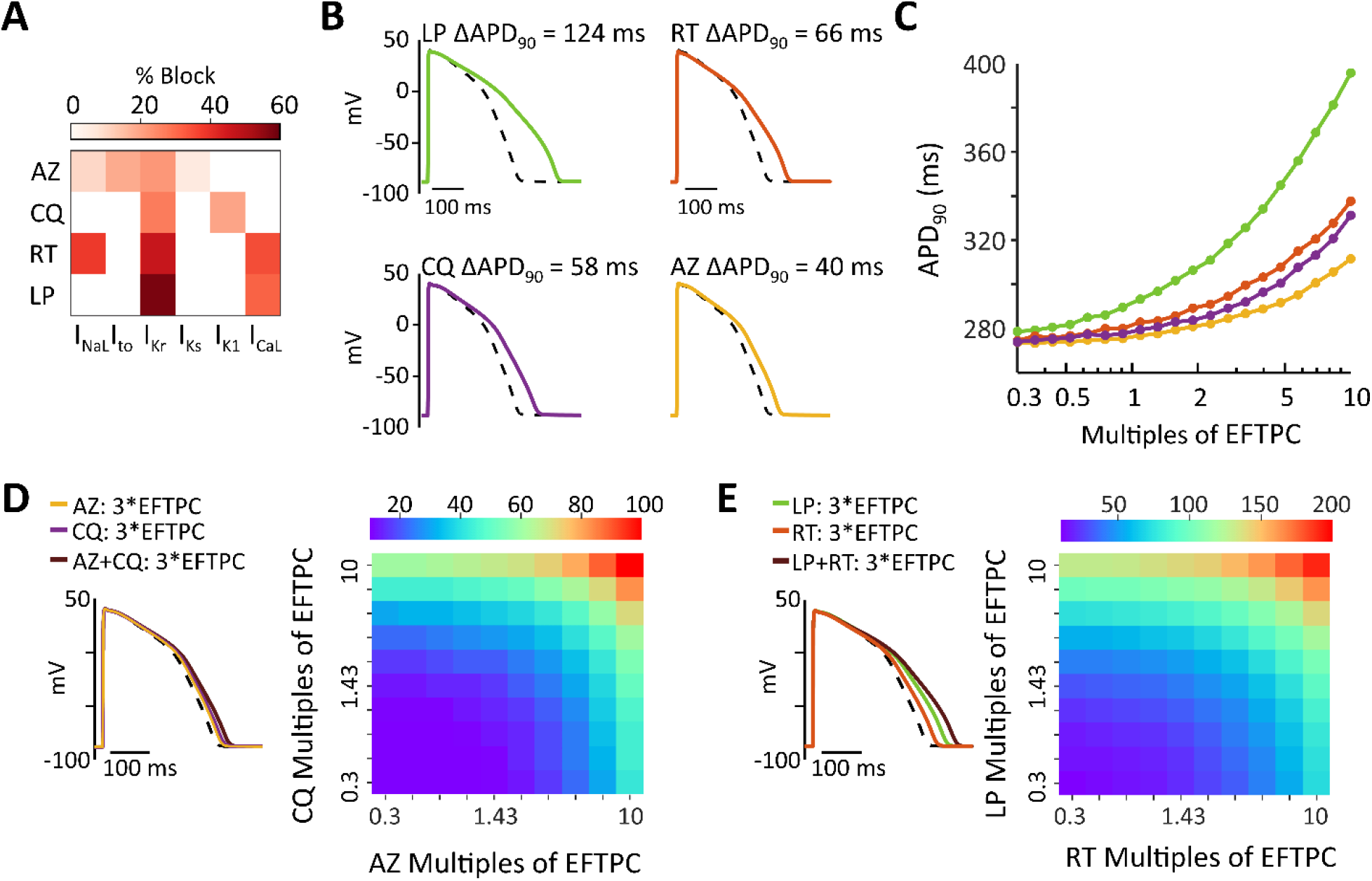
**(A)** Heatmap illustrating the extent to which azithromycin (AZ), chloroquine (CQ), ritonavir (RT), and lopinavir (LP) inhibit 6 important cardiac ionic currents, as previously measured.^12^ Reference 11 reported effective free therapeutic plasma concentration (EFTPC) of each drug, in addition to IC_50_ values that indicated how much each drug influenced 6 cardiac ionic currents (see Methods for abbreviations). Block of currents by particular drugs at 10*EFTPC was calculated based on drug concentration and IC_50_ values using a simple pore block model. **(B)** Simulations with the baseline myocyte model demonstrating how each simulated at 10*EFTPC are predicted to influence ventricular action potentials (APs). **(C)** Concentration-response curves illustrating how the 4 drugs influence APD_90_, the duration between the action potential upstroke (maximal rate of change of voltage) and 90% repolarization. Drug concentrations tested ranged from 0.3 times to 10 times EFTPC, with logarithmically-spaced increments. **(D)** Predicted AP prolongation (∆APD_90_) for chloroquine + azithromycin. **(E)** Predicted ∆APD_90_ for lopinavir + ritonavir. Combination therapy causes greater AP prolongation than drugs applied individually, as shown in both heatmaps illustrating ∆APD_90_ over a range of drug concentrations, and in example AP traces showing effects at 3*EFTPC.

Because recently-published clinical studies on COVID-19 treatments have delivered combination therapy to at least some patients,^1, 2^ we next simulated the combined effects of lopinavir + ritonavir^1^ and chloroquine + azithromycin.^2^ Importantly, the simulations predict that combination therapy causes a greater increase in APD_90_ (ΔAPD_90_) than does either drug in isolation. For instance (**Fig. 1D**), at 3 times EFTPC, lopinavir and ritonavir individually produce ∆APD_90_ of 50.5 and 25.2 ms, respectively, whereas ∆APD_90_ is 73.9 ms for combination therapy. Similarly (**Fig.1E**), ∆APD90 for chloroquine + azithromycin is larger (32.4 ms) than that produced by either drug in isolation (19.8 and 12.6 ms, respectively).

Although the mechanistic simulations indicate the possibility for pharmacodynamic interactions during combination therapy, drug concentrations in patients may not reach the levels assumed in the simulations. To more accurately incorporate clinical drug concentrations, we examined quantitative PK studies on these drugs.^6, 7 8^ Simulations with a model developed for lopinavir + ritonavir therapy,^7^ shown in Fig 2A, predict higher lopinavir concentrations with combination therapy (400 mg/100 mg twice daily) than with lopinavir therapy alone (400 mg twice daily). This occurs due to PK interactions whereby ritonavir inhibits lopinavir clearance. Simulations were next performed in a virtual population to predict how drug plasma concentrations vary between individuals. These results suggest that, between extreme individuals in a population, peak concentrations of either drug may differ by more than 10-fold. Importantly, the results predict that plasma concentrations for lopinavir (Fig. 2B) approach IC_50_ values for cardiac ionic currents in some individuals whereas plasma concentrations of ritonavir (Fig. 2C) remain considerably lower than IC_50_ values. Predictions of chloroquine PK were less straightforward to simulate, given uncertainty in the literature about both protein binding^15^ and accumulation of drug in target tissues, including the heart.^6^ For chloroquine + azithromycin combination therapy, we therefore combined predictions of a PK model for azithromycin^8^ (daily dosing, 500 mg) with published concentrations from a physiologically-based PK model for chloroquine that incorporated drug accumulation in different tissues.^6^ These simulations (Fig. 2D) predict that azithromycin concentrations generally remain well below reported cardiac ionic current IC_50_ levels whereas chloroquine concentrations in the heart can exceed IC_50_ values and block ion channels, due to drug accumulation. To link the predictions of the PK models with QSP model simulations, we calculated the mean peak concentration of the 5% of the patients with the highest drug concentrations (highlighted in red in Fig. 2) and used those values as an input.

**Figure 2:**
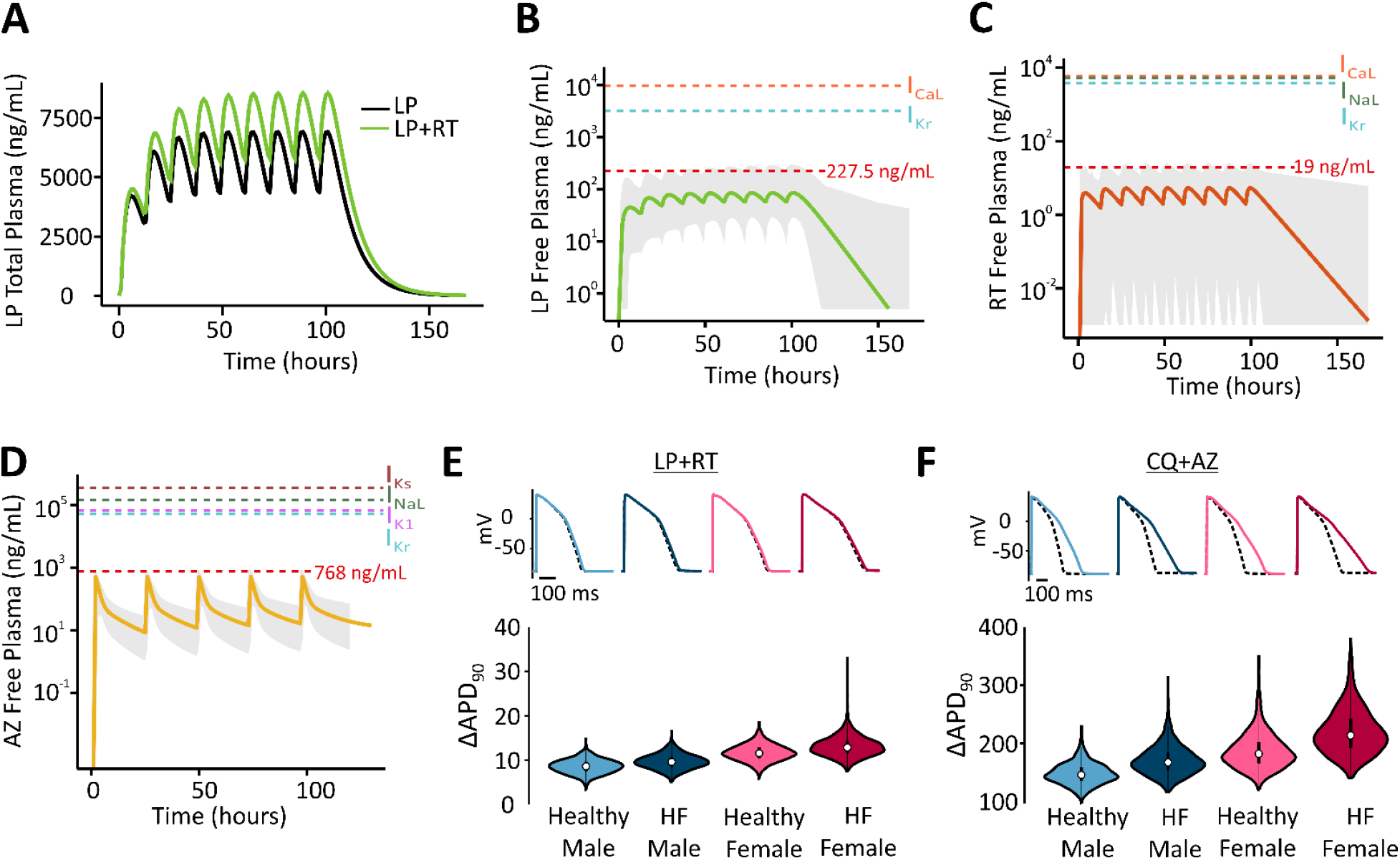
**(A)** Simulations show that plasma concentrations of lopinavir are greater with coadministration of ritonavir than when the former drug is given alone. **(B)** Predicted free plasma concentrations of lopinavir under standard dosing regimen with ritonavir, for median individual (thick line) within a virtual population of 1000 individuals (gray shaded area). Red dashed line indicates mean peak concentration of the 5% of the patients with highest drug concentrations. Additional dashed lines indicate IC_50_ values for cardiac ion channel inhibition. **(C)** Predicted free plasma concentrations of ritonavir under standard dosing regimen with lopinavir, displayed as in **(B)**. **(D)** Predicted free plasma concentrations of azithromycin, displayed as in **(B)**. **(E)** Distributions of ∆APD_90_ caused by clinical concentrations of lopinavir + ritonavir, in 4 virtual patient populations, as indicated. **(F)** Distributions of ∆APD_90_ caused by clinical concentrations of chloroquine + azithromycin, in 4 virtual patient populations, as indicated. The free chloroquine concentration used as an input was 3900 ng/ml based on the original publication results (total chloroquine concentrations in the heart around 10,000 ng/ml multiplied by the unbound fraction of 39%).

Finally we performed simulations to predict how sex differences and pre-existing cardiac disease may influence the AP prolongation caused by COVID-19 therapies. To do this, we created virtual populations of ventricular myocytes representing 4 patient groups: healthy males, healthy females, males with heart failure, and females with heart failure. Simulations of both lopinavir + ritonavir (Fig. 2E) and chloroquine + azithromycin (Fig. 2F), using drug concentrations from the PK simulations, predict that AP prolongation is greatest in the female heart failure group. For lopinavir + ritonavir therapy, median AP prolongation is below 20 ms in all groups, but can reach as high as 30 ms for individual cells in the female heart failure population. Chloroquine + azithromycin therapy, which produced greater AP prolongation in general, caused arrhythmic dynamics in many more cells in the female heart failure population (63 cells from a population of 1000) compared with other groups (1 cell in male healthy, 13 cells in male heart failure, 8 cells in female healthy).

Overall the simulation results indicate that proposed treatments for COVID-19 do indeed carry cardiac risk, and special caution should be exercised when developing combination therapies. With standard dosing regimens, the simulations suggest that AP prolongation is primarily driven by either lopinavir or chloroquine in the two combination therapy regimens examined, with chloroquine causing the most pronounced effects. This result is consistent with a recent small clinical trial in Brazil, which was terminated due to increased mortality in the high-dose chloroquine arm of the study.^2^ An improved safety profile may be a reason for clinicians to favor the chloroquine derivative hydroxychloroquine for COVID-19,^23^ but this drug is also associated with cardiac adverse events.^3^ The simulations of virtual populations suggest that women with pre-existing heart disease will be most at greatest risk of arrhythmia during treatment with these drugs. This increased vulnerability of women, combined with the generally worse prognosis for men with COVID-19,^24^ suggests that sex differences should be considered when weighing risks against benefits. Overall, our study indicates how simulations of both drug disposition and drug mechanisms can be used to identify patient groups at risk of adverse events, thereby helping to guide treatment decisions during this rapidly-evolving pandemic.

## Data Availability

N/A

## Author Contributions

All authors collaboratively conceptualized the work and designed the research. MV performed simulations of ventricular action potentials, and II-A performed the pharmacokinetics simulations. EAS wrote the manuscript. All authors assisted in editing the manuscript.

